# Quality of antenatal care and perinatal outcomes: evidence from a cohort study in Ethiopia, Kenya, South Africa, and India

**DOI:** 10.1101/2025.04.10.25325357

**Authors:** Wen-Chien Yang, Shalom Sabwa, Anagaw Derseh Mebratie, Beatrice Amboko, Irene Mugenya, Sein Kim, Emily R Smith, Monica Chaudhry, Nokuzola Cynthia Mzolo, Nompumelelo Gloria Mfeka-Nkabinde, Theodros Getachew, Tefera Taddele, Damen Haile Mariam, Sailesh Mohan, Prashant Jarhyan, Margaret E. Kruk, Catherine Arsenault

## Abstract

**Background:** Antenatal care (ANC) is crucial for ensuring the health of pregnant women and their newborns. Although ANC coverage has improved globally, ANC quality remains suboptimal in some settings. Evidence on the association between ANC quality and perinatal outcomes in low-resource countries is still limited. Hence, this study assessed ANC quality and its relationship with fetal loss and low birth weight (LBW) newborns.

**Methods and findings:** We used data from the eCohort study that collected longitudinal data on ANC utilization and quality until the end of pregnancy across eight sites in Ethiopia, Kenya, South Africa, and India. Women were enrolled from public government-owned facilities only in India and South Africa and from both public and private facilities in Ethiopia and Kenya. Primary outcomes included fetal loss (≥13 weeks of gestation) and LBW. Good quality ANC was defined as receiving six essential care components during the first ANC visit: blood pressure measurement, blood and urine tests, ultrasound, iron and folic acid supplementation, and counseling on pregnancy danger signs. We conducted mixed-effect logistic regressions to assess the association between good quality ANC and perinatal outcomes, with a sensitivity analysis where good quality ANC excluded ultrasound scans. Among 3,597 pregnant women followed until the end of pregnancy, only 5.8% received all six essential care components during their first ANC visit (ranging from 1.4% in India to 14.0% in Ethiopia) and 30.7% received five care components (excluding ultrasounds) ranging from 5.7% in India to 52.5% in Kenya. Fetal loss prevalence was 3.7% in Ethiopia, 3.8% in Kenya, 4.0% in South Africa, and 6.0% in India. India and South Africa had higher rates of LBW newborns (among neonates who were alive at the time of the survey): 16.3% and 13.1%, respectively, compared to 8.6% in Ethiopia and 8.5% in Kenya. Multiple pregnancies were rarely detected at the first ANC visit. Good quality ANC was associated with a 22% to 58% lower risk of fetal loss (RR 0.78, 95% CI 0.61 – 0.95 to RR 0.42, 95% CI 0.10 – 0.73). No statistically significant associations were observed between good quality ANC and LBW.

**Conclusions:** This study identified important gaps in ANC quality and found that receiving essential ANC services was associated with a lower risk of fetal loss. With increasing global ANC coverage, future research should continue assessing quality, and programs should prioritize quality improvement, ensuring the delivery of good clinical practice and proven evidence-based interventions in pregnancy.

**AUTHOR SUMMARY:** *Why was this study done?:*

- ANC utilization has significantly increased in low-resource settings; however, the quality of care received remains insufficient.
- Most prior research investigating the associations between ANC and perinatal outcomes has focused on ANC utilization or the number of visits.
- While the global dialogue has gradually shifted to ANC quality rather than merely ANC utilization, limited evidence has explored the association between ANC quality and critical perinatal outcomes, including fetal loss and LBW newborns.

*What did the researchers do and find?:*

- We used data from a longitudinal study that collected information on the utilization and quality of ANC throughout pregnancy and assessed the care received and its relationship with fetal and neonatal outcomes.
- Our study found that ANC quality was overall poor. Only 6% of pregnant women received all six care components (blood pressure measurement, blood and urine tests, ultrasound examination, iron and folic acid given or prescribed, and counseling on pregnancy danger signs) at their first ANC visits and only 31% received five care components (excluding ultrasounds).
- Good quality ANC was significantly associated with a lower risk of fetal loss (including late miscarriage and stillbirth), while no significant association was found between the receipt of good quality ANC and LBW newborns.

*What do these findings mean?:*

- Poor-quality antenatal care is not only inefficient but can also be harmful. This issue is particularly concerning as increasing numbers of women in low-resource settings seek ANC services.
- Efforts at the national level should ensure the delivery of good clinical practice and essential care components for high quality ANC to improve perinatal outcomes.

## INTRODUCTION

Antenatal care (ANC) is crucial for ensuring the health of pregnant women and their babies. ANC serves critical functions, including health promotion, screening and diagnoses of diseases, management of maternal morbidities and pregnancy complications, and prevention of illnesses throughout pregnancy.[1] The 2016 World Health Organization (WHO) recommendations on ANC emphasize the importance of high-quality ANC in delivering the right care at the right time.[1]

Previous research has linked ANC to reduced adverse neonatal outcomes. However, a large body of evidence has primarily focused on the utilization of ANC, or in other words, the number of ANC visits. Two meta-analyses concluded that at least one ANC visit reduced the risk of neonatal mortality by 34% to 39% in Asia and sub-Saharan Africa.[2, 3] A study using 193 Demographic and Health Surveys (DHS) from 69 countries showed that at least one ANC visit was associated with reduced neonatal and infant mortality.[4] Studies have also demonstrated that ANC utilization has protective effects against fetal loss and low birth weight (LBW).[4–9] Given the benefits of ANC, low- and middle-income countries (LMICs), where most maternal and newborn mortality and morbidities occur, have made tremendous efforts to increase ANC coverage. However, quality of care remains insufficient, even for women who initiate ANC early in their pregnancy and meet the recommended number of visits.[10–12]

Evidence on the relationship between ANC quality and fetal and neonatal outcomes—such as stillbirths and LBW newborns—remains relatively limited compared to its association with other neonatal outcomes, including neonatal death. A 2022 UNICEF report on stillbirths in LMICs indicated notable evidence gaps in estimating the overall effect of ANC quality on stillbirths.[13] Nearly two million babies are stillborn every year, 98% in LMICs, causing significant economic and psychosocial consequences.[14–16] Antepartum stillbirths are linked to several modifiable risk factors, including maternal infections, non-communicable diseases, poor nutrition, and lifestyle factors. [15, 17–20] Hence, high quality ANC is crucial for addressing these risk factors. Few studies have assessed associations between ANC quality and stillbirths. One study from Ghana found an association between good quality ANC and a reduced risk of stillbirth [8]; another study in Nairobi, Kenya, also demonstrated the protective effects of good quality ANC against stillbirth.[7] However, both studies have design limitations: the former used data from a cross-sectional population-based survey, which may be prone to recall bias and limitations in causal inference, while the latter retrospectively reviewed medical records with a small sample size.

Although the prevalence has declined slowly over the past two decades, LBW remains a significant global health concern, with an estimated prevalence of 14.7% in 2020, affecting approximately 19.8 million newborns worldwide.[21] LBW newborns are not only at a significantly higher risk of morbidity and mortality but also more likely to develop non-communicable diseases, such as hypertension and diabetes, in adulthood.[22, 23] The recently introduced “small vulnerable newborns” framework integrates preterm infants, small-for-gestational-age newborns, and those with LBW, advocating for a comprehensive and integrative approach to addressing the burden of small newborns.[24, 25] ANC should incorporate evidence-based interventions and essential care components to prevent and manage LBW. Some research suggested that ANC attendance and good quality ANC reduced LBW.[4–6, 9]

Critical evidence gaps remain in understanding the association between ANC quality and perinatal outcomes in low-resource settings. Hence, this study aimed to describe ANC quality, identify key care components of good quality ANC and estimate its association with fetal loss and the risk of LBW in four LMICs: Ethiopia, Kenya, South Africa, and India.

## METHODS

### Data source

We used data from the Maternal and Newborn Health (MNH) eCohort study, a longitudinal mixed-mode survey (face-to-face and phone surveys) on maternal and newborn health care quality in four countries (Ethiopia, Kenya, South Africa, and India). The study was conducted in two sites in each country, one rural and one urban. Pregnant women were selected from health facilities and enrolled after their first ANC visit. The health facilities selected included only public government-owned facilities in India and South Africa and both public and private facilities in Ethiopia and Kenya. All women seeking ANC for the first time in their pregnancy, regardless of gestational age, and planning to carry the pregnancy to term were eligible for inclusion. Details of the MNH eCohort study design and methodology, including a description of study sites and recruitment, are available elsewhere.[26–28] The enrollment surveys conducted between April 2023 and January 2024 included in-person health assessments and a review of maternal health cards. The follow-up surveys were conducted via phone approximately every 4 weeks during pregnancy, and after delivery or end of pregnancy was reported. Surveys covered the number of ANC visits, the content of care received, the participant’s health, and the newborn’s health if the pregnancy resulted in a live birth. The analytic sample for the present study included participants who were followed until the end of pregnancy and excluded women who were lost to follow-up as well as those who reported early miscarriages (pregnancy loss before 13 weeks of gestation) since miscarriages in the first trimester are generally caused by chromosomal abnormalities and are rarely preventable.[29, 30]

### Measures

We included two primary outcomes: fetal loss and LBW newborns. Fetal loss was defined as any pregnancy loss after 13 weeks of gestation, including late miscarriages and stillbirths. Late miscarriages were defined as pregnancy losses between 13 and 28 weeks of gestation, and stillbirths were defined as pregnancy losses after 28 weeks of gestation (excluding those born alive) in accordance with the WHO case definition.[31] Newborns’ birth weights were self-reported by mothers. For women who did not know the actual birth weight, we used the survey question of the Demographic and Health Survey that asked women to recall the baby’s birth size, including whether the baby was “very large, larger than average, average, smaller than average, or very small” at birth. Our primary case definition of LBW included newborns with birth weights less than 2.5 kg and newborns described as “smaller than average” or “very small” if actual birth weight data was unavailable. Data on birth body weight or mother-reported birth body size were only collected for newborns who were still alive at the time of the final follow-up phone survey. Gestational age (GA) at baseline, which was used to classify late miscarriages or stillbirths, was estimated based on the date of last menstrual period (LMP) reported by the woman, the estimated due date (EDD) provided by the health worker, or according to the self-reported weeks of pregnancy if the former two dates were unknown or unavailable in the health records. GA was further categorized into trimesters, with the first trimester defined as <13 weeks of gestation, the second trimester defined as ≥13 to <28 weeks, and the third trimester defined as ≥28 weeks.

### Antenatal care quality

The primary independent variable was good quality ANC, a binary variable indicating the receipt of all six basic care components at the first ANC visit: blood pressure (BP) was measured, blood test performed (either a blood draw or finger prick), urine test performed, ultrasound examination, iron and folic acid (IFA) given or prescribed, and counseling on pregnancy danger signs. These six essential care components were chosen based on WHO recommendations and previous studies on ANC quality.[1, 5, 7, 8, 32, 33] We also created a good quality ANC binary variable based on the receipt of five care components (excluding ultrasound scan). We chose to exclude ultrasound scans because the South African ANC guidelines at the time of the study did not recommend them for routine care. In addition, WHO recommends at least one ultrasound scan before 24 weeks of gestation. Therefore, women initiating ANC early might receive the ultrasound in subsequent visits before 24 weeks of gestation. Although the eCohort study obtained information on the ANC content of care for follow-up visits, we restricted the care components to those received at the first visit to avoid immortal time bias since pregnant women who experience fetal loss after the first visit cannot obtain additional ANC. Including data from subsequent visits would bias the association upward and overestimate the protective effect of ANC.[34]

### Covariates

We included independent variables potentially associated with the primary outcomes, including maternal age, marital status, whether the pregnancy was intended, education level (categorized into three groups: no education or some primary education, complete primary education, and complete secondary education or higher), health literacy (defined as answering six health knowledge questions correctly), and household wealth (based on ownership of certain assets and categorized into country-specific wealth tertiles).[27] Second, we included a series of covariates related to the pregnant woman’s health at baseline: self-rated health (very good or excellent, compared to good, fair, and poor), reporting any danger signs in pregnancy at baseline (vaginal bleeding, fever, fainting or loss of consciousness), and having risk factors at baseline (categorized by the number of risk factors: no risk factor, one risk factor, two risk factors, and three or more risk factors). The risk factors assessed included any chronic illness(es) known before the pregnancy, a history of obstetric complications (including Cesarean sections, preterm birth, stillbirth, neonatal death, or postpartum hemorrhage), HIV, and multiple pregnancy detected at the first ANC visit.

### Statistical analysis

First, we present descriptive characteristics of pregnant women at baseline and the proportion of women who received all six and all five and each of the six care components in each country. Second, we reported the prevalence of fetal and neonatal outcomes (late miscarriage, stillbirth, and LBW). Third, we conducted mixed-effect logistic regressions to investigate the association between good quality ANC and fetal loss and LBW, respectively. Regressions included country-fixed effects, two-level random intercepts (for the study site and the health facility where the woman attended ANC), and robust standard errors. We used post-estimation commands and marginal probabilities to obtain risk ratios for the association between good quality ANC and each primary outcome.

Regressions were performed at the fetal and neonatal level. The first model assessed associations between good quality ANC and fetal losses (late miscarriages or stillbirths). The fetal loss regression was restricted to women who were enrolled in their first trimester of pregnancy to avoid selection bias. Including pregnant women who were already in the second or third trimester at baseline would introduce bias since pregnancies that have progressed to the second or third trimester are less likely to develop late miscarriages (which by definition can only occur in the second trimester between 13 and 28 weeks)). In other words, participants who would have passed the period during which particular outcomes of interest could occur should be excluded from the sample. The second regression model assessed associations between good quality ANC and LBW. The LBW regression model included all women (regardless of pregnancy stage at enrollment) but controlled for GA (in trimester) at the first ANC visit.

We performed two sensitivity analyses. First, for fetal losses, we restricted the sample to women with more reliable baseline GA, defined as a baseline GA calculated based on LMP or EDD, and excluded women whose baseline GA was self-reported based on the number of weeks they thought to be pregnant. Second, we conducted a sensitivity analysis for LBW newborns that restricted the sample to those newborns with actual birth weight data and excluded those with birth body sizes described as “smaller than average or very small.” Each regression model (including sensitivity analyses) was repeated using the independent variable of good quality ANC with five components that excluded ultrasounds. Statistical significance was determined based on a p-value < 0.05, and 95% confidence intervals were provided. All analyses were performed using STATA version 18.0.

### Ethical approval

The study protocol was reviewed and approved by the Institutional Review Boards (IRB) of the Harvard T.H. Chan School of Public Health (protocol #IRB22-0487), the Kenya Medical Research Institute (protocol number KEMRI/SERU/CGMR-C/4226), the Ethiopian Public Health Institute (protocol number EPHI-IRB-448-2022), the University of KwaZulu-Natal (protocol number BREC/00004645/2022), and the Public Health Foundation of India (protocol number TRC-IEC 495/22). Formal informed consent was obtained from all adult study participants or emancipated minors or from formal guardians or next-of-kins for study participants under the age of 18 years old in accordance with local regulations.

## RESULTS

### Characteristics of study participants

This study included 3,597 women followed from their first ANC visit until the end of pregnancy **(Table 1)**. The mean age of participants was 25.9 years old (range 24.5 to 26.9), with more adolescents enrolled in South Africa. Nearly all participants in Ethiopia (97.7%) and India (99.9%) and the majority in Kenya (78.1%) were married or partnered, compared to a small proportion in South Africa (11.7%). Pregnancy intention varied widely, with 90.9% in India, 73.4% in Ethiopia, 60.9% in Kenya, and only 16.9% in South Africa reporting the current pregnancy as intended. Maternal health status at baseline varied. Around 60% of women in India (66.7%) and Kenya (56.0%) rated their health as very good or excellent, compared to about 40% in South Africa and Ethiopia. In India, 19.2% of women had at least one risk factor, followed by 24.8% in Ethiopia and 28.0% in Kenya; in contrast, this proportion was markedly higher in South Africa at 50.2%, where HIV prevalence was the highest (28.8%). Few women in India (5.5%) and Kenya (8.3%) reported any pregnancy danger signs at baseline, compared to 13.1% in Ethiopia and 25.6% in South Africa. Half of the women in India initiated ANC visits in the first trimester, compared to only 13.8% to 38.0% in the three sub-Saharan African countries. In addition, few women were detected with multiple pregnancies during their first ANC visit. In Ethiopia, Kenya, and South Africa, only 43.8% (7/16), 22.2% (4/18), and 26.3% (5/19) of multiple pregnancies identified at delivery were detected at the first ANC visit, respectively. However, in India, three twin pregnancies detected at the first ANC visit were singleton at delivery.

**Table 1.** Characteristics of pregnant women followed until the end of pregnancy included in the study by country^1^.

|  | Overall<br>N=3597<br>Frequency (%) | Ethiopia<br>N=883<br>Frequency (%) | Kenya<br>N=896<br>Frequency (%) | South Africa<br>N=854<br>Frequency (%) | India<br>N=964<br>Frequency (%) |
| --- | --- | --- | --- | --- | --- |
| **Sociodemographic** |  |  |  |  |  |
| Age, years (mean/standard deviation) | 25.9 (0.1) | 25.6 (0.2) | 26.9 (0.2) | 26.7 (0.2) | 24.5 (0.1) |
| Age category, years |  |  |  |  |  |
| 15 to <20 | 389 (10.8%) | 73 (8.3%) | 104 (11.6%) | 142 (16.6%) | 70 (7.3%) |
| ≥20 to <35 | 2885 (80.2%) | 753 (85.3%) | 667 (74.4%) | 591 (69.2%) | 874 (90.7%) |
| ≥35 | 323 (9.0%) | 57 (6.5%) | 125 (14.0%) | 121 (14.2%) | 20 (2.1%) |
| Married or partnered | 2625 (73.1%) | 862 (97.7%) | 700 (78.5%) | 100 (11.7%) | 963 (99.9%) |
| Intended pregnancy | 2211 (61.8%) | 648 (73.4%) | 546 (60.9%) | 144 (16.9%) | 873 (90.9%) |
| Education level |  |  |  |  |  |
| No education or some primary | 557 (15.5%) | 334 (37.8%) | 65 (7.3%) | 6 (0.7%) | 152 (15.8%) |
| Complete primary | 1221 (34.0%) | 353 (40.0%) | 313 (34.9%) | 248 (29.1%) | 307 (31.9%) |
| Complete secondary or higher | 1817 (50.5%) | 196 (22.2%) | 518 (57.8%) | 599 (70.2%) | 504 (52.3%) |
| Health literacy<sup>2</sup> | 1032 (28.7%) | 237 (26.8%) | 398 (44.4%) | 277 (32.4%) | 120 (12.5%) |
| Wealth status<sup>3</sup> |  |  |  |  |  |
| Poorest | 1127 (31.6%) | 283 (32.7%) | 299 (33.6%) | 228 (26.7%) | 317 (33.1%) |
| Middle | 1395 (39.1%) | 286 (33.1%) | 383 (43.0%) | 339 (39.7%) | 387 (40.4%) |
| Richest | 1044 (29.3%) | 296 (34.2%) | 209 (23.5%) | 286 (33.5%) | 253 (26.4%) |
| **Maternal health** |  |  |  |  |  |
| Self-rated own health as very good or excellent<sup>4</sup> | 1842 (51.2%) | 335 (37.9%) | 502 (56.0%) | 362 (42.4%) | 643 (66.7%) |
| Risk factor at baseline<sup>5</sup> |  |  |  |  |  |
| No risk factor | 2510 (69.8%) | 664 (75.2%) | 643 (72.0%) | 425 (49.8%) | 779 (80.8%) |
| One risk factor | 797 (22.2%) | 184 (20.8%) | 192 (21.4%) | 280 (32.8%) | 142 (14.7%) |
| Two risk factors | 199 (5.5%) | 29 (3.3%) | 33 (3.7%) | 104 (12.2%) | 33 (3.4%) |
| Three or more risk factors | 91 (2.5%) | 6 (0.7%) | 28 (3.1%) | 45 (5.3%) | 10 (1.0%) |
| HIV status<sup>6</sup> | 277 (7.7%) | 13 (1.5%) | 15 (1.7%) | 244 (28.8%) | 5 (0.5%) |
| **Pregnancy characteristics** |  |  |  |  |  |
| Trimester of the first ANC visit |  |  |  |  |  |
| First trimester<sup>7</sup> | 1187 (33.3%) | 259 (29.9%) | 124 (13.8%) | 319 (38.0%) | 485 (50.3%) |
| Second trimester<sup>7</sup> | 2045 (57.4%) | 559 (64.6%) | 613 (68.4%) | 444 (52.9%) | 429 (44.5%) |
| Third trimester<sup>7</sup> | 334 (9.4%) | 48 (5.5%) | 159 (17.8%) | 77 (9.2%) | 50 (5.2%) |
| Report at least one danger sign at baseline<sup>8</sup> | 461 (12.8%) | 116 (13.1%) | 74 (8.3%) | 218 (25.6%) | 53 (5.5%) |
| Multiple pregnancy known at baseline | 24 (0.7%) | 7 (0.8%) | 4 (0.5%) | 5 (0.6%) | 8 (0.8%) |
| Multiple pregnancy identified at delivery | 59 (1.6%) | 16 (1.8%) | 18 (2.0%) | 19 (2.2%) | 6 (0.6%) |

<sup>1</sup> Participants who had an early miscarriage (fetal loss before 13 weeks of gestation) or were lost to follow-up after the first ANC visit were excluded.
<sup>2</sup> Defined as answering six health knowledge questions correctly. The six questions were adopted from the Indian Health and Human Development Survey.
<sup>3</sup> Wealth tertile is country-specific.
<sup>4</sup> Self-rated own health was based on a Likert scale of five levels (excellent, very good, good, fair, and poor).
<sup>5</sup> Includes any chronic systemic illness(es) known before pregnancy (including diabetes, hypertension, cardiac disease, HIV, mental health disorder, schizophrenia, epilepsy, seizure, renal disorder, asthma, tuberculosis, anemia, hemoglobinopathy, chronic pelvic inflammatory disease, ovarian cyst, fibroids, uterine myoma, genital tract abnormalities, thyroid cancer, thyroid disease, peptic ulcer disease, gestational hypertension in previous pregnancy, a history of stroke) and any history of obstetric complications (Cesarean section, stillbirth, preterm birth, neonatal death, and postpartum hemorrhage)
<sup>6</sup> HIV was reported separately in this table while it was included in the development of categorization of risk factors.
<sup>7</sup> The first trimester is defined as less than 13 weeks of gestation; the second trimester is defined as between 13 to less than 28 weeks of gestation; the third trimester is defined as above 28 weeks of gestation
<sup>8</sup> Includes vaginal bleeding, fever, and fainting or loss of consciousness.

### Antenatal care quality

Only 5.8% of women received all six care components at their first ANC visit, ranging from 1.4% only in India to 14.0% in Ethiopia (**Table 2**). Use of ultrasound was low, ranging from only 7.6% of women in South Africa to 43.4% in Ethiopia. Counseling on pregnancy danger signs was highest in Kenya (60.9%), followed by 49.4% in South Africa, 31.0% in Ethiopia, and lowest in India (15.0%). Iron and folic acid supplements were given or prescribed to most women, from 80.8% in Ethiopia to 93.1% in Kenya. Excluding ultrasounds increased the proportion of women who received all five care components to 30.7%. Results stratified by research sites and by facility ownership in each country are shown in **Supplemental Table 1 and Supplemental Table 2**. When excluding private facilities, ANC quality ranged from 1.4% in India to 6.9% in Kenya.

**Table 2.** Proportion of pregnant women who received each of the six and all six care components at first ANC visits by country.

| ANC quality care components | Overall<br>N=3597 | Ethiopia<br>N=883 | Kenya<br>N=896 | South Africa<br>N=854 | India<br>N= 964 |
| --- | --- | --- | --- | --- | --- |
|  | Frequency (%) | Frequency (%) | Frequency (%) | Frequency (%) | Frequency (%) |
| **Physical examination** |  |  |  |  |  |
| Blood pressure measurement | 3304 (91.9%) | 640 (72.5%) | 861 (96.2%) | 852 (99.8%) | 951 (98.8%) |
| **Test and examinations** |  |  |  |  |  |
| Blood test (blood draw or finger prick) | 3419 (95.1%) | 838 (94.9%) | 876 (97.9%) | 853 (99.9%) | 852 (88.5%) |
| Urine test | 3045 (84.7%) | 752 (85.2%) | 801 (89.5%) | 850 (99.5%) | 642 (66.6%) |
| Ultrasound | 709 (19.8%) | 383 (43.4%) | 74 (8.3%) | 65 (7.6%) | 187 (19.5%) |
| **Counseling** |  |  |  |  |  |
| Signs of pregnancy complications | 1380 (38.5%) | 273 (31.0%) | 544 (60.9%) | 420 (49.4%) | 143 (15.0%) |
| **Preventions** |  |  |  |  |  |
| Iron and folic acid given or prescribed | 3217 (89.4%) | 713 (80.8%) | 831 (93.1%) | 790 (92.7%) | 883 (92.2%) |
| **Completeness of all six care components<sup>1</sup>** | 209 (5.8%) | 124 (14.0%) | 50 (5.6%) | 22 (2.6%) | 13 (1.4%) |
| **Completeness of all five care components<sup>2</sup>** | 1105 (30.7%) | 179 (20.3%) | 470 (52.5%) | 401 (47.0%) | 55 (5.7%) |

<sup>1</sup> Include blood pressure measurement, blood test, urine test, ultrasound examination, iron and folic acid given or prescribed, counseling on signs of pregnancy complications
<sup>2</sup> Include all the care components except for ultrasound examination.

### Perinatal outcomes

**Table 3** describes fetal and neonatal outcomes. The proportion of late miscarriages ranged from 2.3% in Ethiopia to 3.9% in India, while stillbirths ranged from 1.3% to 2.1%. Among neonates who were alive at the post-delivery follow-up survey, India had the highest rates of LBW infants (16.3%), compared to 8.5% to 13.1% in the other three countries. Data on EDD, LMP, and actual newborn birth weight was available for the majority of the observations in India, Kenya, and South Africa. However, in Ethiopia, data on EDD or LMP was available for only half (51.1%) of neonates, and actual newborn birth weight data was available for 59.6% of newborns.

**Table 3.** Fetal and neonatal outcomes.

|  | **Overall**<br>**N=3657** | **Ethiopia**<br>**N=900** | **Kenya**<br>**N=914** | **South Africa**<br>**N=873** | **India**<br>**N=970** |
| --- | --- | --- | --- | --- | --- |
| **Fetal outcomes** | **Frequency (%)** | **Frequency (%)** | **Frequency (%)** | **Frequency (%)** | **Frequency (%)** |
| Late miscarriage<sup>1</sup> | 104 (2.8%) | 21 (2.3%) | 21 (2.3%) | 24 (2.8%) | 38 (3.9%) |
| Stillbirth<sup>2</sup> | 57 (1.6%) | 12 (1.3%) | 14 (1.5%) | 11 (1.3%) | 20 (2.1%) |
| Livebirth | 3496 (95.6%) | 867 (96.3%) | 879 (96.2%) | 838 (96.0%) | 912 (94.0%) |
| **Fetal outcomes**<br>**(among those with more reliable baseline gestational age)<sup>3</sup>** | **N=2923 (79.9%)<sup>6</sup>** | **N=460 (51.1%)<sup>6</sup>** | **N=875 (95.7%)<sup>6</sup>** | **N=868 (99.4%)<sup>6</sup>** | **N= 720 (74.2%)<sup>6</sup>** |
| Late miscarriage<sup>1</sup> | 80 (2.7%) | 9 (2.0%) | 20 (2.3%) | 24 (2.8%) | 27 (3.8%) |
| Stillbirth<sup>2</sup> | 46 (1.6%) | 6 (1.3%) | 13 (1.5%) | 11 (1.3%) | 16 (2.2%) |
| **Neonatal outcomes**<br>**(among neonates still alive at the post-delivery survey)** | **N=3441<sup>7</sup>** | **N=851<sup>7</sup>** | **N=862<sup>7</sup>** | **N=828<sup>7</sup>** | **N=900** |
| Low birth weight newborn<sup>4</sup> | 398 (11.7%) | 72 (8.6%) | 73 (8.5%) | 106 (13.1%) | 147 (16.3%) |
| **Neonatal outcomes**<br>**(among neonates still alive at the post-delivery survey and who**<br>**had data on actual birth weights)** | **N=3021 (87.8%)<sup>8</sup>** | **N=507 (59.6%)<sup>8</sup>** | **N=842 (97.7%)<sup>8</sup>** | **N=773 (93.4%)<sup>8</sup>** | **N=899 (99.9%)<sup>8</sup>** |
| Low birth weight newborn<sup>5</sup> | 346 (11.5%) | 37 (7.3%) | 69 (8.2%) | 93 (12.0%) | 147 (16.4%) |

<sup>1</sup> Defined as fetal loss between 13 to 28 weeks of gestation
<sup>2</sup> Defined as fetal loss after 28 weeks of gestation
<sup>3</sup> More reliable baseline gestational age was determined by last menstrual period or estimated due date.
<sup>4</sup> Defined as newborn birth body weight less than 2.5kg, or self-reported newborn size as smaller than average or very small if actual birth weight was unavailable.
<sup>5</sup> Defined as newborn birth body weight less than 2.5kg.
<sup>6</sup> The percentage was calculated by dividing the number with a more reliable baseline gestational age (using last menstrual period to determine) by the number with a more reliable baseline gestational age (using last menstrual period to determine) or mothers' self-reported weeks of pregnancy if last menstrual period and estimated due date were unknown.
<sup>7</sup> The percentage was calculated by dividing the number with actual birth weight by the number with actual birth weight and mothers' self-reported birth size if actual birth body weight was unknown.
<sup>8</sup> Among neonates who were still live at the post delivery survey, 13 (out of 851) in Ethiopia, 4 (out of 862) in Kenya, and 18 (out of 828) in South Africa had missing data for both birth weight and size.

### Results of mixed-effect logistic regressions

Risk ratios (RR) for the association between good quality ANC and perinatal outcomes from the eight regression models are summarized in **Figure 1**. Receiving all six ANC components was associated with a 58% reduced risk of fetal loss (late miscarriages or stillbirths, RR 0.42, 95% CI 0.10–0.73). In models with good quality ANC that excluded ultrasounds, both primary and sensitivity analyses also showed statistically significant associations between good quality ANC and fetal loss (RR 0.68, 95% CI 0.45-0.92 and RR 0.78 95% CI 0.61, 0.95). Regressions for LBW newborns, both primary and sensitivity analyses, did not show statistically significant associations between receiving good quality ANC and LBW newborns.

**Figure 1.**
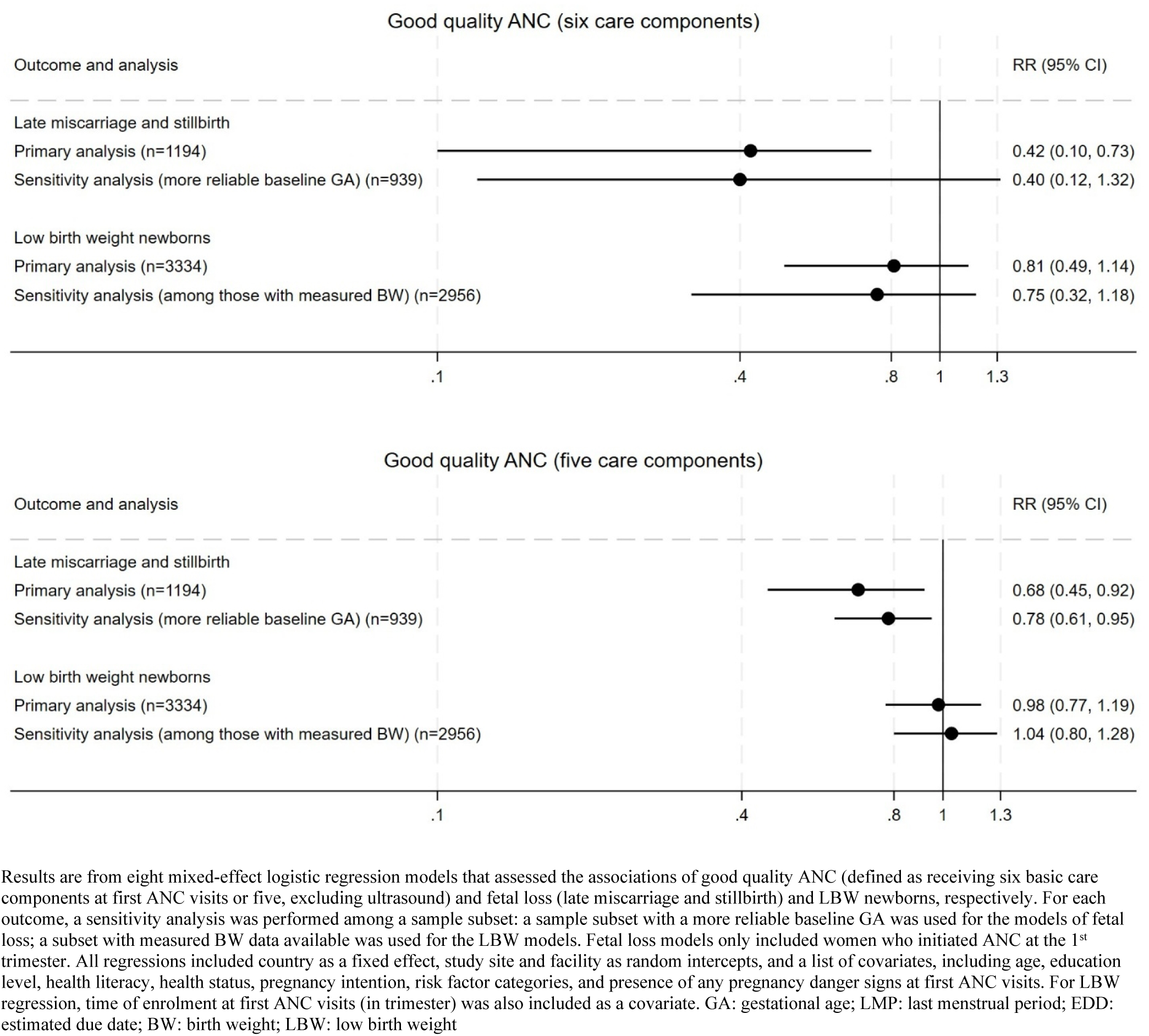
Risk ratios (RR) of fetal loss (late miscarriage and stillbirth) and LBW newborns between those who received and who did not receive good quality ANC.

In addition, we found that pregnant women who reported any pregnancy danger signs at baseline had increased risks of miscarriages or stillbirths in both regression models where good quality ANC had six components (Odds ratio (OR) 1.89, 95% CI 1.24–2.86) and five components (OR 1.94, 95% CI 1.28–2.95) **(Table 4)**. In the regressions for LBW newborns, mothers giving birth in South Africa and India were more likely to have a LBW newborn compared to those in Ethiopia. Women aged 20 to 35 years old, compared to those younger than 20 years old, had significantly lower odds of delivering LBW newborns in both models where good quality ANC had six or five components (OR 0.80, 95% CI 0.69 – 0.93; OR 0.80, 95% CI 0.69 – 0.94) **(Supplemental Table 3)**.

**Table 4.** Results of mixed effect logistic regressions for fetal loss (late miscarriage and stillbirth)

|  | Good quality ANC<br>(received six care components)<br>N = 1194 |  |  | Good quality ANC<br>(received five care components)<br>N= 1194 |  |  |
| --- | --- | --- | --- | --- | --- | --- |
|  | Odds<br>Ratio<br>(OR) | 95%<br>Confidence<br>Interval (CI) | p-value | Odds<br>Ratio<br>(OR) | 95%<br>Confidence<br>Interval (CI) | p-value |
| **Received good quality ANC** | **0.39** | **0.18 – 0.87** | **0.022** | **0.66** | **0.45 – 0.97** | **0.036** |
| **Socioeconomic and demographic status** |  |  |  |  |  |  |
| *Country* |  |  |  |  |  |  |
| Ethiopia | *ref* |  |  | *ref* |  |  |
| Kenya | 0.81 | 0.47 – 1.41 | 0.466 | 0.98 | 0.49 – 1.97 | 0.964 |
| South Africa | 0.50 | 0.17 – 1.52 | 0.223 | 0.61 | 0.18 – 2.05 | 0.423 |
| India | 1.17 | 0.66 – 2.08 | 0.594 | 1.21 | 0.65 – 2.23 | 0.551 |
| *Age category, years* |  |  |  |  |  |  |
| 15 to <20 | *ref* |  |  | *ref* |  |  |
| ≥20 to <35 | 0.77 | 0.49 – 1.21 | 0.257 | 0.77 | 0.49 – 1.21 | 0.264 |
| ≥35 | 1.59 | 0.47 – 5.40 | 0.460 | 1.58 | 0.46 – 5.45 | 0.472 |
| *Intended pregnancy* | 0.89 | 0.48 – 1.66 | 0.715 | 0.89 | 0.47 – 1.68 | 0.718 |
| *Education level* |  |  |  |  |  |  |
| No education or some primary | *ref* |  |  | *ref* |  |  |
| Complete primary | 1.10 | 0.67 – 1.78 | 0.713 | 1.07 | 0.65 – 1.77 | 0.787 |
| Complete secondary or higher | 1.03 | 0.54 – 1.97 | 0.921 | 1.02 | 0.53 – 1.95 | 0.958 |
| *Health literacy<sup>1</sup>* | 1.57 | 0.91 – 2.74 | 0.108 | 1.59 | 0.92 – 2.74 | 0.098 |
| *Wealth* |  |  |  |  |  |  |
| Poorest | *ref* |  |  | *ref* |  |  |
| Middle | 0.89 | 0.57 – 1.38 | 0.602 | 0.90 | 0.59 – 1.39 | 0.638 |
| Richest | 0.85 | 0.67 – 1.08 | 0.195 | 0.86 | 0.69 – 1.07 | 0.178 |
| **Maternal health** |  |  |  |  |  |  |
| *Self-rated own health as very good or excellent<sup>2</sup>* | 1.10 | 0.60 – 2.00 | 0.764 | 1.11 | 0.61 – 1.99 | 0.739 |
| *Risk factor at baseline<sup>3</sup>* |  |  |  |  |  |  |
| No risk factor | *ref* |  |  | *ref* |  |  |
| One risk factor | 1.41 | 0.87 – 2.28 | 0.164 | 1.43 | 0.89 – 2.31 | 0.141 |
| Two risk factors | 1.48 | 0.64 – 3.42 | 0.354 | 1.45 | 0.66 – 3.18 | 0.359 |
| Three or more risk factors | 2.46 | 0.94 – 6.44 | 0.067 | **2.65** | **1.05 – 6.69** | **0.039** |
| *Reported any pregnancy danger sign at baseline<sup>4</sup>* | **1.89** | **1.24 – 2.86** | **0.003** | **1.94** | **1.28 – 2.95** | **0.002** |

<sup>1</sup> Defined as answering six health knowledge questions correctly. The six questions were adopted from the Indian Health and Human Development Survey.
<sup>2</sup> Self-rated own health was based on a Likert scale of five levels (excellent, very good, good, fair, and poor)
<sup>3</sup> Risk factors at baseline include any chronic systemic illness(es) known before pregnancy (including diabetes, hypertension, cardiac disease, HIV, mental health disorder, schizophrenia, epilepsy, seizure, renal disorder, asthma, tuberculosis, anemia, hemoglobinopathy, chronic pelvic inflammatory disease, ovarian cyst, fibroids, uterine myoma, genital tract abnormalities, thyroid cancer, thyroid disease, peptic ulcer disease, gestational hypertension in previous pregnancy, a history of stroke), any history of obstetric complications (Cesarean section, stillbirth, preterm birth, neonatal death, and postpartum hemorrhage), and multiple pregnancy known at the first ANC visit
<sup>4</sup> Pregnancy danger signs at baseline include vaginal bleeding, fever, and fainting or loss of consciousness.

## DISCUSSION

In this study, we used data from the MNH eCohort study in four countries to investigate associations between ANC quality and perinatal outcomes. We found that less than 10% of pregnant women in the study received all six key care components at their first ANC visit. While the majority of women had their blood pressure measured and gave blood and urine samples, the proportion of women receiving ultrasound examinations and being counseled on pregnancy danger signs was much lower. Our analysis also showed that good quality ANC (defined by receiving at least five or six care components during the first ANC visit) was associated with a reduced risk of fetal loss but not LBW.

The association between good quality ANC and fetal loss confirms the importance of good quality ANC. Notably, we observed larger effect sizes in models where good quality ANC was defined as receiving six care components compared to models that excluded ultrasounds from defining good quality. This specific finding highlights the value of ultrasound scans. Obstetric ultrasounds, if done early, allow for accurate GA assessment, monitoring fetal growth (if done repeatedly) as well as placenta problems that may be linked to stillbirth, identifying congenital anomalies, and detecting breeched babies and obstructed labor (if used around the time of delivery) – all of which are crucial for the health of pregnant women and their babies.[35, 36] While we found low receipt of ultrasound scans at first visits, we acknowledged that these countries recommend an ultrasound scan before 24 weeks of gestation, which might occur after the first visit. Since 90% of women in our study started ANC visits in the first and second trimesters, many may not receive a scan at the first visit based on national recommendations. Low ultrasound coverage might also explain why multiple pregnancies were rarely detected at the first visit. For example, few women in South Africa had an ultrasound during their first ANC visit, and the majority (74%) of multiple pregnancies were not detected at that visit. This phenomenon is particularly concerning, as multiple pregnancy is a well-established risk factor for numerous adverse outcomes.[37] Early identification of multiple pregnancies by obstetric ultrasound helps risk stratification and timely referrals. There remain critical gaps in obstetric ultrasound use in LMICs: first, access to ultrasound equipment is still limited [38, 39]; second, obstetric ultrasounds are mainly diagnostic, and the ability to link scan results with management strategies for improving health outcomes varies depending on health provider competence.[40] A 2014 cluster randomized trial assessed the effects of two ultrasound scans in pregnancy, one at 16 to 22 weeks and one at 32 to 36 weeks, on maternal and neonatal mortality in five LMICs (Zambia, Kenya, the Democratic Republic of the Congo, Pakistan, and Guatemala). The study showed an anticipated higher detection of multiple pregnancies but no improvement in maternal, fetal, and neonatal survival.[40] In our study, assessing early ultrasound scans in the first ANC visit, an aspect overlooked in prior research, offers critical insights.[1]

Our work contributes to the growing evidence on the benefits of ANC quality in the context of increasing ANC coverage.[10–12, 32, 41] In a study using household surveys from 91 LMICs, only two-thirds of women who used ANC received three essential care components (BP measurement, blood test, and urine test).[12] Many pregnant women in low-resource countries who receive the recommended number of ANC visits do not receive essential care content.[10, 11] A study across 10 LMICs found that while most women in need of ANC had at least one visit, only two-fifths attended four or more. In addition, the quality of care was suboptimal, with blood pressure measurement being the most commonly performed service, while counseling on complications was the least provided.[10] A population-based study in India found poor coverage of quality-adjusted ANC, defined as receiving BP and weight measurements, abdomen examinations, and blood and urine tests.[32] Our results reinforce existing evidence on the insufficient quality of ANC, urging global efforts to prioritize the delivery of necessary care and evidence-based interventions at ANC visits rather than merely increasing the number of ANC contacts. In addition to poor quality ANC, another interesting finding is the significant association between self-reported danger signs and fetal loss at baseline. Future research should explore the quality of ongoing management of danger signs that may lead to severe pregnancy complications.

In our study, between 8.5% to 16.3% of babies were LBW. However, the actual proportion of LBW in our sample may be higher, as birth weight or size data were unavailable for neonatal and infant deaths. In addition, there was a higher prevalence of LBW in South Africa, which might be partly attributable to higher prevalences of adolescent pregnancies and maternal HIV infection. LBW babies, along with preterm and SGA–viewed as a large population of “small vulnerable newborns”–are not only at an increased risk for mortality throughout the first year of life but also at a higher risk for suboptimal neurodevelopment.[42, 43] Many evidence-based antenatal nutritional interventions have been proven to prevent LBW, such as IFA, balanced protein/energy, and multiple micronutrient supplementation.[44, 45] Our study included IFA in defining good quality ANC and explored its relationship with LBW, but no associations were found. There are a few potential explanations. First, the care components we identified were proxies of good quality ANC, which might not capture all essential care components that affect LBW newborns. Future studies might consider including other care content and investigating how it relates to LBW. Second, receiving the care components does not guarantee adherence to medical advice or appropriate management when risk factors are detected.[46]

The present study had limitations. First, our analyses and inference testing are limited by small sample sizes. Late miscarriages and stillbirths are rare outcomes; only 4.4% of women in our sample experienced them, leading to large standard errors. Similarly, some sensitivity analyses were likely underpowered due to small sample sizes. Second, our results may have limited generalizability as the study enrolled participants from two particular sites in each country, not nationally representative samples. Third, we only included care components at the first ANC visit to define good quality. These components, while fundamental, are surrogate indicators for the overall ANC quality throughout pregnancy. The rationale for not including care content at follow-up visits was to avoid immortal time bias, a systematic bias that occurs when the exposure definition includes a period during which the outcome cannot occur, potentially leading to spurious associations. In perinatal epidemiology studies, this bias might significantly affect the analyses examining the effects of pregnancy exposures on stillbirths or miscarriages.[34, 47] In our study, we did not include follow-up visits because women who experience a miscarriage or stillbirth after the first ANC visit cannot receive additional antenatal care, and thus the follow-up period is no longer comparable. Fourth, newborn birth weight and size data were only collected for babies alive at the post-delivery follow-up survey, excluding the birth weight data for neonatal or infant deaths. As LBW babies are at a higher risk of death compared to normal-weight babies, some cases of neonatal and infant deaths likely had low birth weights but were excluded from our study. Fifth, the LBW analysis was limited by the mother’s self-reported baby birth size, with only 60% of babies in Ethiopia having actual birth weight data, compared to 90% in the other three countries. In addition, we were unable to distinguish between antepartum and intrapartum stillbirths in the fetal loss analysis. Lastly, these countries differed in their national ANC guidelines. For instance, Ethiopia and Kenya recommend an ultrasound scan for all women before 24 weeks of gestation, while South Africa’s 4^th^ edition national ANC guidelines recommend an ultrasound scan only for women who are unsure about their LMP (and the 5^th^ edition guidelines have changed to recommend a scan for everyone). This might explain the low receipt of ultrasound scans in South Africa.

Delivering high-quality ANC remains a challenge in LMIC settings with the highest burdens of maternal and newborn morbidity and mortality. Future research should not only assess overall ANC quality and its impact on perinatal outcomes but also focus on the quality of risk detection and the ability of health providers to manage preexisting and emerging risk factors in pregnancy effectively. Meanwhile, policies should prioritize the implementation of good clinical practices and the essential components that define high-quality ANC.

## Data Availability

The analytic dataset will be available from Harvard Dataverse.

## ADDITIONAL INFORMATION

### Supporting information captions

Supplemental Table 1. Proportion of pregnant women who received each of the six and all six care components at first ANC visits in four countries by research sites

Supplemental Table 2. Proportion of pregnant women who received each of the six and all six care components at first ANC visits in four countries by facility ownership

Supplemental Table 3. Results of mixed effect logistic regressions for low birth weight newborns

### Author contributions

#### Conceptualization

Wen-Chien Yang, Catherine Arsenault, and Margaret E. Kruk

#### Funding

Margaret E. Kruk

#### Formal analysis

Wen-Chien Yang, Catherine Arsenault

#### Writing

Wen-Chien Yang and Catherine Arsenault wrote the first draft. Shalom Sabwa, Anagaw Derseh Mebratie, Beatrice Amboko, Irene Mugenya, Sein Kim, Emily R Smith, Monica Chaudhry, Nokuzola Cynthia Mzolo, Nompumelelo Gloria Mfeka-Nkabinde, Theodros Getachew, Tefera Taddele, Damen Haile Mariam, Sailesh Mohan, Prashant Jarhyan, Margaret E. Kruk, and Catherine Arsenault critically reviewed the manuscript, made edits, provided feedback on the interpretations and suggested relevant literature.

